# Preliminary evidence that higher temperatures are associated with lower incidence of COVID-19, for cases reported globally up to 29th February 2020

**DOI:** 10.1101/2020.03.18.20036731

**Authors:** Melanie Bannister-Tyrrell, Anne Meyer, Céline Faverjon, Angus Cameron

**Affiliations:** Ausvet, 5 Shuffrey Street, Fremantle, Western Australia, Australia; Ausvet Europe, 3 Rue Camille Jordan, Lyon, France

**Keywords:** COVID-19, SARS-CoV-2, coronavirus, pandemic, temperature, seasonality

## Abstract

Seasonal variation in COVID-19 incidence could impact the trajectory of the pandemic. Using global line-list data on COVID-19 cases reported until 29^th^ February 2020 and global gridded temperature data, and after adjusting for surveillance capacity and time since first imported case, higher average temperature was strongly associated with lower COVID-19 incidence for temperatures of 1°C and higher. However, temperature explained a relatively modest amount of the total variation in COVID-19 incidence. These preliminary findings support stringent containment efforts in Europe and elsewhere.

## Background

Pandemic COVID-19, caused by a beta-coronavirus named SARS-CoV-2 first identified in Wuhan, China [1], has spread rapidly, particularly in temperate regions in the northern hemisphere [2]. The number of cases reported in countries in tropical regions, most of which are low- and middle-income countries (LMICs) with weaker detection and response capacity [3], is lower [2]. There has been much speculation about whether warming temperatures with the onset of spring and summer will contribute to decreased transmission in the northern hemisphere, as is observed for many viral respiratory infections [4]. This would also have implications for the risk of spread in temperate regions in the southern hemisphere at the onset of winter, and for tropical regions where the vast majority of LMICs are located. Higher temperatures were shown to have a protective effect against transmission of severe acute respiratory syndrome (SARS) in 2002–2003 [5], possibly due to the decreased survival of the SARS-CoV on surfaces at higher temperatures [6]. Decreased aerosol spread at higher temperatures is another possible mechanism, as observed for human influenza viruses [7]. This study aimed to provide preliminary data on whether there may be seasonal variation in COVID-19 incidence, in sub-national regions that have reported at least one confirmed COVID-19 case.

## Methods

An open-source line list of confirmed COVID-19 cases was downloaded on March 2^nd^ 2020 [8]. The line list included data on confirmed cases up to February 29^th^ 2020 for all countries, including China. Cases were aggregated to the first-level administrative division (ADM1) in which they occurred, as defined by the Global Administrative Areas dataset (https://gadm.org/, accessed March 4^th^ 2020). This corresponds to the first-level administrative unit within each country, usually described as a state or province. The reported coordinates of the case (variably a point location, city centroid, or different subnational administrative levels) were used to determine the ADM1 in which the case occurred. For each ADM1, an observation period was defined as period from the date of onset of symptoms of the first reported case for that ADM1 to February 29^th^ 2020. When information regarding onset of symptoms of the first reported case was missing, the confirmation date of the first reported case was used instead.

Daily gridded temperature data at 0.5-degree spatial resolution were obtained from the Climate Prediction Centre (NOAA/OAR/ESRL PSD, Boulder, Colorado, USA, https://www.esrl.noaa.gov/psd/, accessed March 4^th^ 2020). The average temperature at the ADM1 centroid was calculated by taking the average of the maximum and minimum temperatures over the observation period at the centroid coordinates, using packages {ncdf4} [9] and {rgdal} [10]. All the analyses were implemented in the R environment [11].

All ADM1 associated with at least one confirmed case of COVID-19 in the source dataset were included in the analysis, excluding Hubei province in China where the outbreak emerged. Each case was classified as imported (using the proxy that travel history was reported in the line list data) or local (otherwise). We modelled the cumulative number of COVID-19 cases classified as local cases in each ADM1 during the observation period. The statistical model was based on the generalized linear regression framework [12]. We used a negative binomial distribution to account for the overdispersion of the number of local cases during the observation period. The primary exposure was the average temperature at the ADM1 centroid during the observation period. Temperature was included in the model both as a linear and a quadratic term to account for the non-linearity of the association observed during initial data exploration.

Four variables were included in the model as potential confounders: the cumulative number of imported cases, time since the first reported case (to account for right-censoring), the median age of the national population (United Nations database, https://ourworldindata.org/age-structure, to account for the higher incidence of severe cases in older people, which may be more readily detected), and the capacity of the country to detect an emerging infectious disease. The Global Health Security Index (GHSI) (https://www.ghsindex.org/) publishes a country-level score (out of 100) for capacity for “early detection and reporting for epidemics of potential concern”. This indicator is a weighted average of indicators related to laboratory systems, real-time surveillance and reporting, epidemiology workforce, and data integration between human, animal and environmental health sectors, each of which reflects a core capacity defined under the International Health Regulations (2005).

Likelihood ratio tests were used to identify variables that did not significantly improve model fit and obtain a final model specification. The number of imported cases was dropped from the final model due to collinearity with the time since first reported case. The final model included time since the first reported case, early detection score and temperature terms. Model fit indicators (leverage, influence, residuals and fitted values) were assessed graphically. Pseudo R-squared values were calculated using the Nagelkerke’s method [13].

## Results and discussion

As of February 29^th^ 2020, 188 ADM1 units worldwide reported at least one imported case and were included as observations in the model. This included 39 provinces in China as well as 149 ADM1-level reports in 46 other countries (Figure 1). A total of 13,479 confirmed cases were included in the dataset.

**Figure 1:**
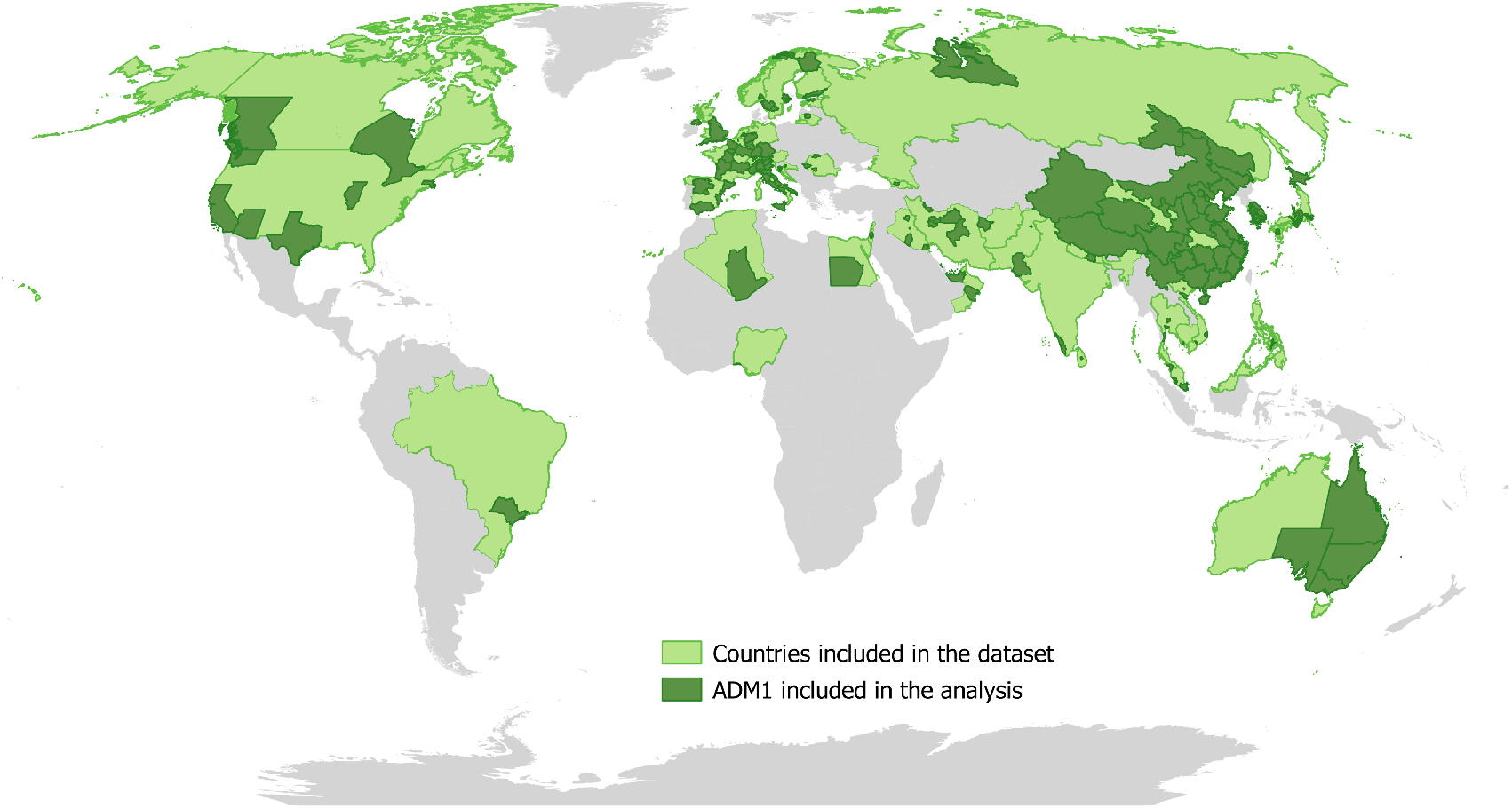
Distribution of ADM1 units that reported at least COVID-19 case up to 29th February 2020.

As of 29^th^ February 2020, provinces in China (excluding Hubei) reported between 0 and 68 imported cases; ADM1 outside China reported between 0 and 26 imported cases (noting that as case travel history was not always reported, classification of imported versus local case was not always possible). The number of presumed locally-transmitted cases ranged from 0 to 1,220 in provinces in China (excluding Hubei), and 0 to 552 in ADM1 outside China (Table 1). Average temperatures at ADM1 centroids in China (excluding Hubei) ranged from -16.8°C to 20.4°C, and from -18.7°C to 31.9°C outside China (Table 1).

**Table 1:** Summary statistics for the variables used in the analysis of the effect of average temperature on the number of local COVID-19 cases at state or province level (N=188 ADM1 units, data as of February 29th, 2020).

| Variable | Type of observation | Min | 25<sup>th</sup> percentile | Median | Mean | 75<sup>th</sup> percentile | Max | s.d. |
| --- | --- | --- | --- | --- | --- | --- | --- | --- |
| Number of imported cases | China (excl. Hubei) | 0 | 1 | 4 | 11 | 14 | 68 | 15 |
|  | Outside China | 0 | 0 | 1 | 2 | 2 | 26 | 3 |
| Number of local cases | China (excl. Hubei) | 0 | 15 | 116 | 257 | 316 | 1,220 | 333 |
|  | Outside China | 0 | 0 | 1 | 12 | 2 | 552 | 58 |
| Average temperature (°C) | China (excl. Hubei) | -16.8 | -0.8 | 2.1 | 2.1 | 6.9 | 20.4 | 8.3 |
|  | Outside China | -18.7 | 4.9 | 8.7 | 10.4 | 16.1 | 31.9 | 10.5 |
| Early detection GHSI score (country-level) | All | 12 | 49 | 70 | 66 | 82 | 98 | 19 |
| Time since first reported case (in days) | China (excl. Hubei) | 1 | 25 | 31 | 29 | 37 | 51 | 14 |
|  | Outside China | 1 | 1 | 3 | 10 | 14 | 56 | 13 |

Average temperature (as a linear and quadratic term combination) was strongly associated with count of local COVID-19 cases (likelihood ratio test = 19.4, df = 2, p=0.00006). Although one observation (Daegu province in South Korea) was identified as an outlier and had high leverage, the model results did not change when this observation was removed from the dataset. The model results indicate that there was a negative correlation in the predicted number of cases with temperature from 1°C and above (Figure 2). For example, at mean values for the other variables, an increase in average temperature from 1°C to 9°C was associated with a decrease in predicted cases at ADM1 level from 24 cases to 19 cases, respectively. Similarly, an increase in average temperature from 10°C to 19°C was associated with a decrease from 18 to 7 predicted cases at ADM1 level, respectively. The pseudo R-squared values for the final model with and without temperature were 0.44 and 0.39, respectively, indicating that the inclusion of the temperature effect only provided a relatively modest improvement in model fit.

**Figure 2:**
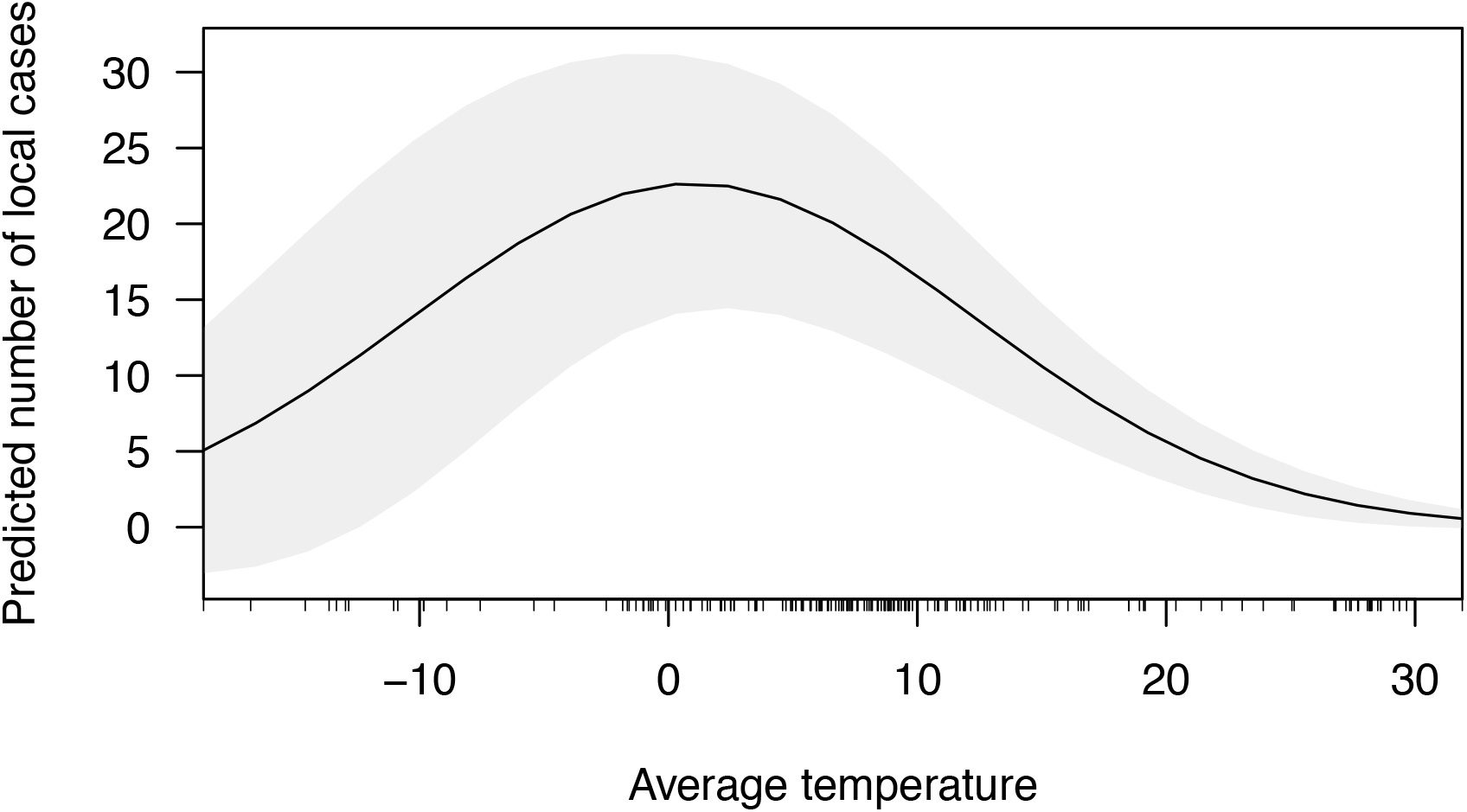
Predicted number of local cases of COVID-19 by 1st-level administrative unit as a function of the average temperature during the period from the first reported case until 29th February 2020. The grey area represents the 95% confidence interval of the predicted values.

There are several important limitations to this study. These results remain preliminary, as they only include confirmed cases as of February 29^th^ 2020, at which point reported local transmission outside China was relatively limited. There was no data available on many characteristics that affect rate of spread within a region, especially the interventions initiated in response to the detection of imported or locally transmitted cases. Furthermore, the model could not be fitted with a random intercept for country to account for clustering of ADM1 units within countries, as the uneven distribution of the number of affected provinces by country led to model instability. However, the early detection score likely captures part of the country-level variance. Is it also important to note that the classification of cases as local or imported was based on available information, and in some ADM1 (in particular in China, South Korea and Italy) the first imported case could not be identified. These results need to be confirmed by repeating the analysis as the pandemic progresses, and including data on implemented interventions to contain or mitigate COVID-19 as it becomes available.

Many LMICs had not detected a COVID-19 case as of 29^th^ February 2020 and therefore were not included in this analysis. Caution is warranted in extrapolating the association between local COVID-19 case counts and temperature to LMICs in tropical regions. COVID-19 outbreaks in LMICs, even if at somewhat lower incidence due to higher temperatures, are still likely to have a substantial impact on health services that are already significantly resource constrained.

This study provides preliminary evidence that there may be seasonal variability in transmission of SARS-CoV-2, but this analysis does not imply that temperature alone is a primary driver of COVID-19 transmission. The observed association may not be due directly to temperature, but to correlated factors such as relative humidity, or human behaviours during cold weather. The findings present an argument for further significant scaling up of containment measures now. The onset of warmer weather in the northern hemisphere may modestly reduce rate of spread, but anticipation of a substantial decline in total number infected due to warmer temperatures alone is not warranted by these findings. Furthermore, implementing stringent containment measures coupled with a seasonal decline in incidence may reduce the risk of COVID-19 becoming endemic in the northern hemisphere, and globally, if transmission in the southern hemisphere can be contained over the same period.

## Data Availability

The study makes use of publicly available line-list data.

https://tinyurl.com/s6gsq5y

https://gadm.org

https://www.esrl.noaa.gov/psd

https://ourworldindata.org/age-structure

## Ethical statement

The analysis made use of publicly available, anonymised data only. Therefore, no approval to conduct the study from an ethical review board was sought. Nonetheless, the study was conducted in accordance with the Declaration of Helsinki, as revised in 2013.

## References

[1] Huang C, Wang Y, Li X, Ren L, Zhao J, Hu Y, et al. Clinical features of patients infected with 2019 novel coronavirus in Wuhan, China. The Lancet 2020;395:497–506.

[2] World Health Organization. Coronavirus disease 2019 (COVID-19) Situation Report - 50. Geneva: 2020.

[3] Gilbert M, Pullano G, Pinotti F, Valdano E, Poletto C, Boëlle P-Y, et al. Preparedness and vulnerability of African countries against importations of COVID-19: a modelling study. The Lancet 2020.

[4] Johnson HC, Gossner CM, Colzani E, Kinsman J, Alexakis L, Beauté J, et al. Potential scenarios for the progression of a COVID-19 epidemic in the European Union and the European Economic Area, March 2020. Eurosurveillance 2020;25:2000202.

[5] Lin K, Fong DY-T, Zhu B, Karlberg J. Environmental factors on the SARS epidemic: air temperature, passage of time and multiplicative effect of hospital infection. Epidemiol Infect 2006;134:223–30.

[6] Chan KH, Peiris JSM, Lam SY, Poon LLM, Yuen KY, Seto WH. The Effects of Temperature and Relative Humidity on the Viability of the SARS Coronavirus. Adv Virol 2011.

[7] Lowen AC, Mubareka S, Steel J, Palese P. Influenza Virus Transmission Is Dependent on Relative Humidity and Temperature. PLOS Pathog 2007;3:e151.

[8] Xu B, Kraemer MUG, Xu B, Gutierrez B, Mekaru S, Sewalk K, et al. Open access epidemiological data from the COVID-19 outbreak. Lancet Infect Dis 2020;0.

[9] Pierce D. ncdf4: Interface to Unidata netCDF (Version 4 or Earlier) Format Data Files. R package version 1.17. 2019. https://CRAN.R-project.org/package=ncdf4.

[10] Bivand R, Keitt T, Rowlingson B. rgdal: Bindings for the “Geospatial” Data Abstraction Library. R package version 1.4-8. 2019. https://CRAN.R-project.org/package=rgdal.

[11] R Core Team. R: A language and environment for statistical computing. Vienna, Austria: 2019. https://www.R-project.org.

[12] Venables WN, Ripley BD. Modern applied statistics with S. Fourth. New York: Springer; 2002.

[13] Nagelkerke NJD. A note on a general definition of the coefficient of determination. Biometrika 1991;78:691–2.

